# The impact of testing and infection prevention and control strategies on within-hospital transmission dynamics of COVID-19 in English hospitals

**DOI:** 10.1101/2020.05.12.20095562

**Authors:** Stephanie Evans, Emily Agnew, Emilia Vynnycky, Julie Robotham

## Abstract

Nosocomial transmission of SARS-CoV-2 is a key concern and evaluating the effect of testing and infection prevention control strategies is essential for guiding policy in this area. Using a within-hospital SEIR transition model of SARS-CoV-2 in a typical UK hospital, we predict that approximately 20% of infections in inpatients, and 89% of infections in HCWs were due to nosocomial transmission. Placing suspected COVID-19 patients in single rooms or bays has the potential to reduce hospital-acquired infections in patients by up to 80%. Periodic testing of HCWs has a smaller effect on the patient-burden of COVID-19 but would considerably reduce infection in HCWs by as much as 64% and result in only a small proportion of staff absences (approximately 1% per day). This is considerably fewer than currently observed due to suspected COVID-19 and self-isolation.

## Introduction

Nosocomial transmission of SARS-CoV-2, the transmission of the virus within a hospital, is a primary concern in managing spread. While the contribution of nosocomial transmission to the spread of COVID-19 is not yet known, hospital-acquired infections played a key role in disease transmission during outbreaks of previous coronavirus diseases. In Severe Acute Respiratory Syndrome (SARS) outbreaks in 2003, patient-to-HCW transmission was identified as a common route of infection (Chowell et al., 2015; de Wit et al., 2016), and patient-to-patient transmission was highly prominent in outbreaks of Middle East Respiratory Syndrome (MERS) in 2012, 2015, and 2016 (Chowell et al., 2015; Hunter et al., 2016; de Wit et al., 2016). It has been suggested that a possible reason for the high level of patient-to-patient transmission during a MERS outbreak was that MERS-CoV RNA was shown to be present in patients’ rooms 4-5 days after patients died or no longer tested positive for the virus (Bin et al., 2016), suggesting that indirect patient-to-patient transmission commonly occurs in nosocomial MERS-CoV infection.

Since the identification of Severe Acute Respiratory Syndrome Coronavirus 2 (SARS-CoV-2) in December 2019 there have been many reports of infections in healthcare workers (HCWs) and hospitalised patients in both the UK and other countries across the world (Hunter et al., 2020; Keeley et al., 2020; Kluytmans et al., 2020; Wang, 2020; Wang et al., 2020). There is considerable uncertainty around the proportion of these cases that developed as a result of nosocomial transmission, rather than community acquisition of the virus. Hospital inpatients represent an almost closed population where estimates of the disease latency can be used to predict the extent to which patients develop symptoms after hospital admission due to progression of pre-symptomatic disease (community acquired) versus those that become infected through nosocomial transmission. In HCWs, however, classifying the source of infection in this population is more difficult as they migrate between the hospital and community on a daily basis. Some studies have estimated the proportion of nosocomial infections among infected HCWs to be around 44% (Zhou et al., 2020), however others have suggested that the majority of infections in HCWs are acquired in the community (Kluytmans et al., 2020). When contact patterns are known, genomic sequencing of pathogens can be used to build transmission trees to determine the origin and transmission chain of an outbreak (Grad and Lipsitch, 2014; Houlihan et al., 2018), however contact tracing is resource-intensive and imperfect (Daskalaki et al., 2007; Dixon et al., 2015; England et al., 2005).

Quantifying and understanding the role of nosocomial transmission in the perpetuation of an epidemic is important for understanding outbreaks and predicting the efficacy of infection prevention and control (IPC) strategies, including testing of both HCWs and the inpatient population. Mathematical models of infection transmission dynamics, when appropriately parameterised, rely on data that is widely collected and are a useful tool for estimating the source of infection in a hospital (Grassly and Fraser, 2008; Keeling and Danon, 2009), and can subsequently be used to test hypotheses around testing and IPC strategies on reducing transmission.

We present a within-hospital SEIR transmission model of SARS-CoV-2, including patients and HCWs and use this model to quantify the contribution of nosocomial infection to total infection burden and effectiveness of alternative control measures in an English hospital.

## Methods

### Model Description

We developed an SEIR transmission model of COVID-19 infection within a hospital, with disease transmission to, from and between both patients and hospital healthcare workers (HCW). The model includes admission testing of suspected cases and that of symptomatic patients and HCW within the hospital. The hospital is split into a general hospital population, as well as three cohorts for suspected cases, test positive and test negative patients. Upon admission to hospital, suspected COVID-19 patients are placed into the suspected cohort (awaiting test results) and subsequently moved to either the positive or negative cohort upon receiving their test result. All admissions that are not tested are admitted to one of the remaining hospital beds, and undetected COVID-19 cases that enter the general hospital compartment can infect other patients and HCWs. HCWs can become infected in the community and from infected patients in the hospital (including positives in cohorts). Figure 1 is a schematic representation of the model.

**Figure 1:**
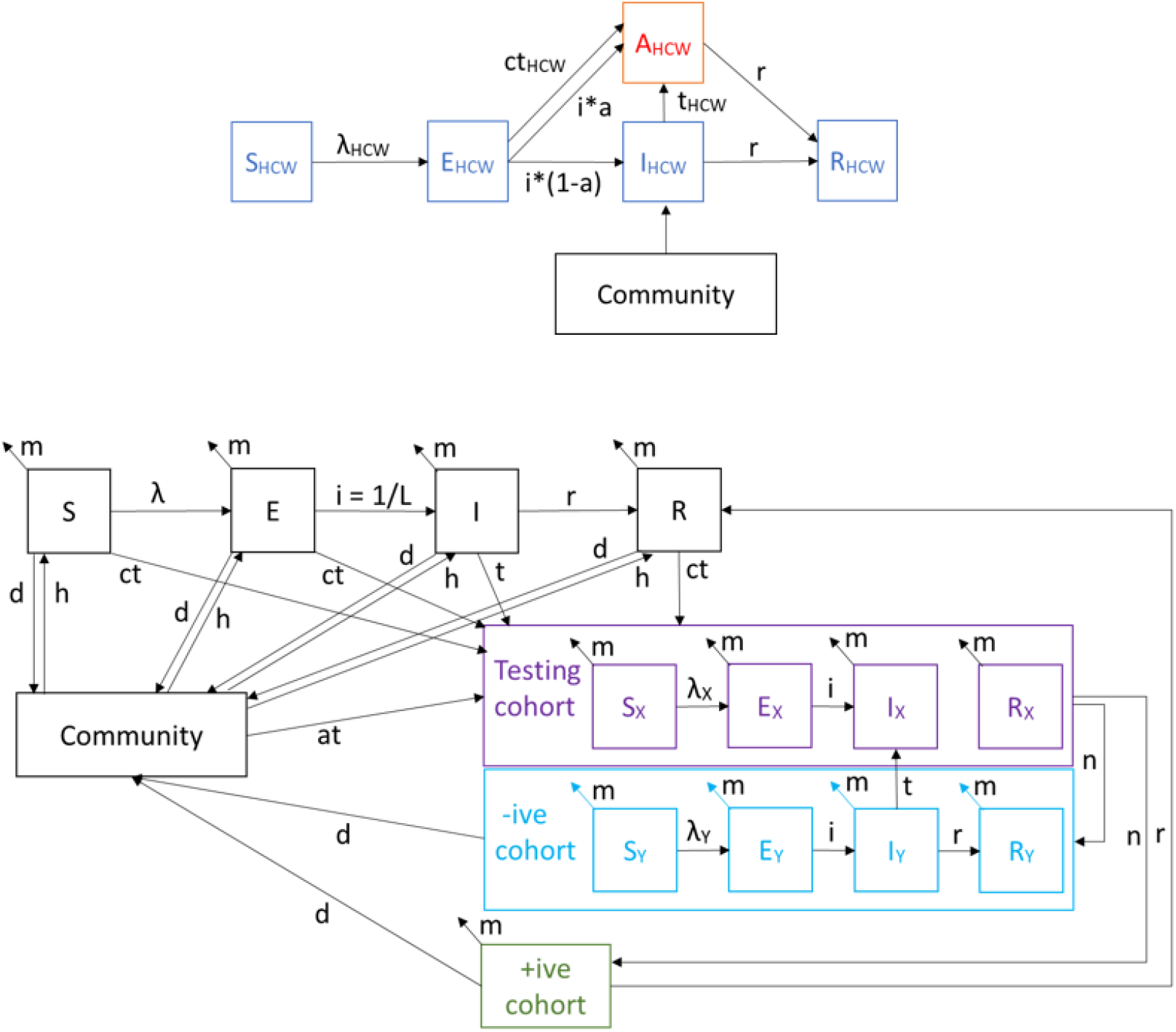
Schematic representation of the model.

Model simulations examine two scenarios for handling suspected cases, each impacting the transmission rates to and from patients to HCWs:

Scenario A: Suspected patients are cohorted together. Amongst those suspected cases will be those who test negative i.e. are uninfected, who have been exposed to cases through being cohorted with those who test positive. In this scenario, transmission between patients in the testing cohort can occur, as well as transmission to and from HCWs.

Scenario B: Suspected patients awaiting test results, are all independently isolated into single-rooms or bays within the hospital, until test results confirm whether to move them to the tested positive or tested negative cohorts. In this scenario, direct transmission between patients in the testing cohort is inhibited, so the parameter for patient to patient transmission is reduced by a factor representative of the effectiveness of single-room cohorting and is varied in a scenario analysis. Patient to HCW transmission is unchanged from Scenario A. The full model code and parameter list can be found in supplementary material.

### Assumptions

This model assumes there are no age, gender or race stratification within patients and HCWs. Beyond the testing and tested cohorts, for which patients cannot directly transmit infection to other patients outside their own cohort, there is no separation of wards within the hospital. The model assumes full immunity of patients and HCWs once infected, no individuals can be reinfected. It also assumes that all exposed patients are pre-symptomatic, none are asymptomatic.

In the tested positive cohort, patients are discharged on recovery and within the suspected positive cohort, it is assumed patients will not be in the cohort long enough to recover from the infection. We have assumed that all patients and HCWs are susceptible to infection at the beginning of the epidemic and that test accuracy is 100%. We also assumed that in the absence of testing 63% of HCW continue to work while infected unless they were tested and found to be positive as observed in a case study in the Netherlands (Kluytmans et al., 2020)

### Parameter Estimation

The model is parameterised for an ‘average’ English hospital, i.e. the admission rate, death rate, and number of cases in hospital reflects the average in England. Parameters were derived from available data where possible, and unknown parameters calibrated to data from the NHS situation report and personal communication with two English Trusts (Table A1).

Following parameter estimation, the proportion of new infections in the community that were admitted each day was maintained across all simulations, but to include a range of infection scenarios, the rates of transmission were varied around the estimated values by multiplying each parameter *p* by a multiplier *m_p_* ~ *N(1,0.25)* (Figure A1).

Parameters and their values, along with the force of infection equations, are provided in the Supplementary File.

### Simulation

Simulations were initialised with a start date of 09-Mar-2020 and ran for 56 simulated days. On initialisation the daily incidence in the community on 09-Mar-2020 was randomly drawn from a Γ (1.42, 57768) distribution to reflect population incidence on day 0 (Figure S1A) and incidence doubled every 2.4 days (Figure S1B) with a cap of 0.0003, the maximum observed daily incidence in the community from PHE’s case tracker (https://www.gov.uk/government/publications/covid-19-track-coronavirus-cases). Infected individuals were imported from the community to the testing cohort at a rate based on the daily incidence in the community at time t - 2 days, since the average time to hospitalisation from onset has been estimated at 2 days (Linton et al., 2020; Pung et al., 2020; Sanche et al.; Tindale et al., 2020; Wang, 2020).

### Uncertainty Analyses

Uncertainty analyses were performed by generating a 1000 random parameter samples and running the model with each parameter set. Partial-rank correlation coefficients were used to calculate the most significant parameter values using the *spartan* R package (Alden et al., 2013).

## Results

### Model fitting and validation

We validated the model against data from the NHS Situation Reports (NHS SitReps). The model was able to replicate the proportion of beds occupied by COVID19 patients longitudinally from NHS Situation Report data (Figure A2). To validate HCW results we compared against PCR swabbing data from two NHS Hospital Trusts (Figure A3).

We performed a sensitivity analysis to identify the most significant parameters of those that were estimated or uncertain (Figure A4). The most significant parameters were the transmission rates between patients and HCWs (bP2P, bH2P, bP2H, bH2H), and the admission rate of asymptomatic and pre-symptomatic cases *(he)*. The probability an exposed HCW would self-isolate (a) was significantly negatively correlated with the proportion of HCWs that were infected.

### Sources of nosocomial transmission to patients and HCWs

We simulated nosocomial infections in patients and HCWs at a typical English hospital from 09-Mar-2020 to 01-May-2020 under Scenario A with the assumption that 37% of symptomatically infected HCW were absent for 7 days following infection, and neither patients nor HCWs could be re-infected. Over the simulated time-period, 13% (IQR 13.2, 14.4) of admitted patients and 11% (IQR 9.1, 14.9) of HCWs were infected with SARS-CoV-2 (Figure 2A,B). Nosocomial transmission was responsible for 20% (IQR 14.4, 27.1) of infections in inpatients, and 89% (IQR 80.2, 93.1) of infections in HCWs, with patients driving nosocomial infections in both cases, although HCW to HCW transmission was also considerable (Figure 2). Patient to patient transmissions were responsible for 25% (IQR 14.1,27.7) of patient infections overall and 78% (IQR 68.7, 87.5) of nosocomial patient infections, and patient to HCW transmissions were responsible for 52% (IQR 43.2,60.4) of all HCW infections and 57% (IQR 46.4, 68.5) of nosocomial HCW infections with HCW to HCW infections responsible for a further 39% of all HCW infections (Figure 2C,D).

**Figure 2:**
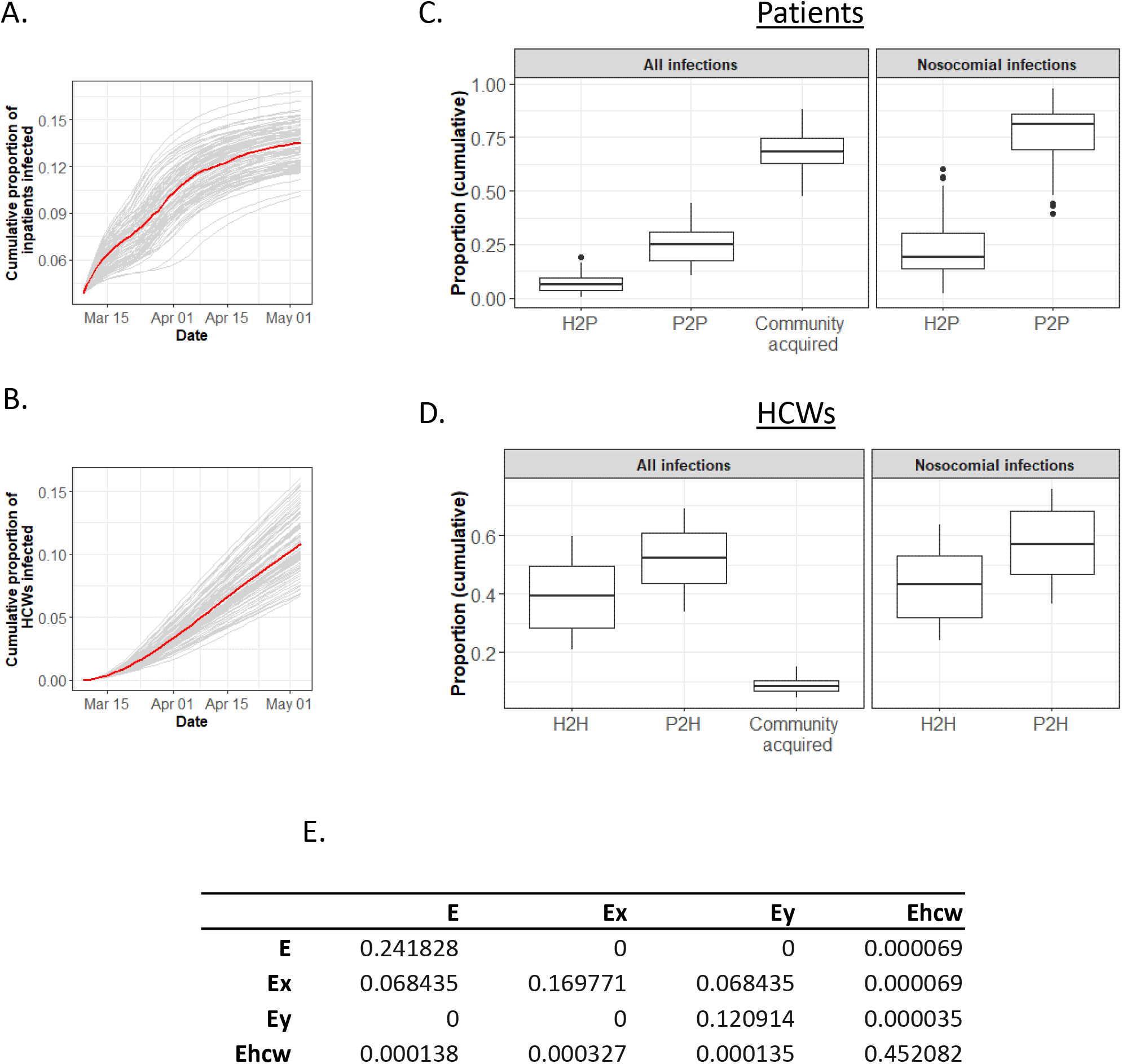
Infection within hospital between 09-03-2020 - 01-05-2020 under Scenario A. A) Cumulative proportion of inpatients that have been infected with COVID-19 (grey = individual simulations (100 reps), red = median). B) Daily cumulative proportion of HCWs that have been infected with COVID19 (grey = individual simulations (100 reps), red = median). C) Total proportion of all infected patients that have had community acquired, hospital acquired infection from direct HCW to patient (H2P), or direct and indirect patient to patient (P2P) transmission by 01-05-2020 (Left panel), and proportion of nosocomial infections attributable to each source (Right panel) D) Total proportion of all infected HCWs that have had community acquired, hospital acquired infection from direct and indirect HCW to HCW (H2W), or direct and indirect patient to HCW (P2H) transmission by 01-05-2020 (Left panel), and proportion of nosocomial infections attributable to each source (Right panel). E) Next-Generation Matrix for infections within hospital, used to estimate within hospital R0. Where, E = Hospitalised people, Ex = those suspected to have infection, Ey = those who have tested negative, and Ehcw = health care workers

We calculated R_0_ within the hospital, and for each source population using the Next Generation Matrix (NGM) approach of Diekmann, Heesterbeek and Roberts (Diekmann et al., 2010) (full details in supplementary material). Using this approach the number of secondary infections among patients resulting from a typical HCW was 1.7*10^−4^, secondary cases in HCWs from each HCW was 0.452, secondary infections in HCWs from a typical patient approximately 0.000138 (given that 91% of exposed asymptomatic patients are on a general ward), and in patients from each patient 0.31 (if suspected patients make up a small proportion of the population) (Figure 2E). The overall within hospital R_0_ is estimated to be 0.45 (IQR 0.36, 0.56).

### Efficacy of periodic testing of HCWs on nosocomial transmission events

Periodic testing of HCWs is a potential tool to prevent the spread of infections in hospital. We considered periodic testing of the entire HCW population every 1, 7, or 14 days under the assumption that following a positive test result, infected and exposed HCWs were absent for 7 days. Daily testing was the most effective at reducing transmission with a reduction of 65% (IQR 57.8, 60.3) in HCW to HCW, and 14% (IQR 12.1,15.9) in HCW to patient transmission events (Figure 3A,B). In the scenario with 7 day periodic testing, infections from HCWs to other HCWs were reduced by 24% (IQR 19.2, 26.7), and periodic testing every 14 days saw a minor effect compared to no testing, 6% (IQR 4.2,7.3) of HCW to HCW transmission events prevented. Under these testing strategies, a maximum of 1.5% of HCWs were absent due to being infected with COVID-19 on any one day, suggesting the workforce is not significantly reduced by the implementation of stringent testing strategies (Figure 3C).

**Figure 3:**
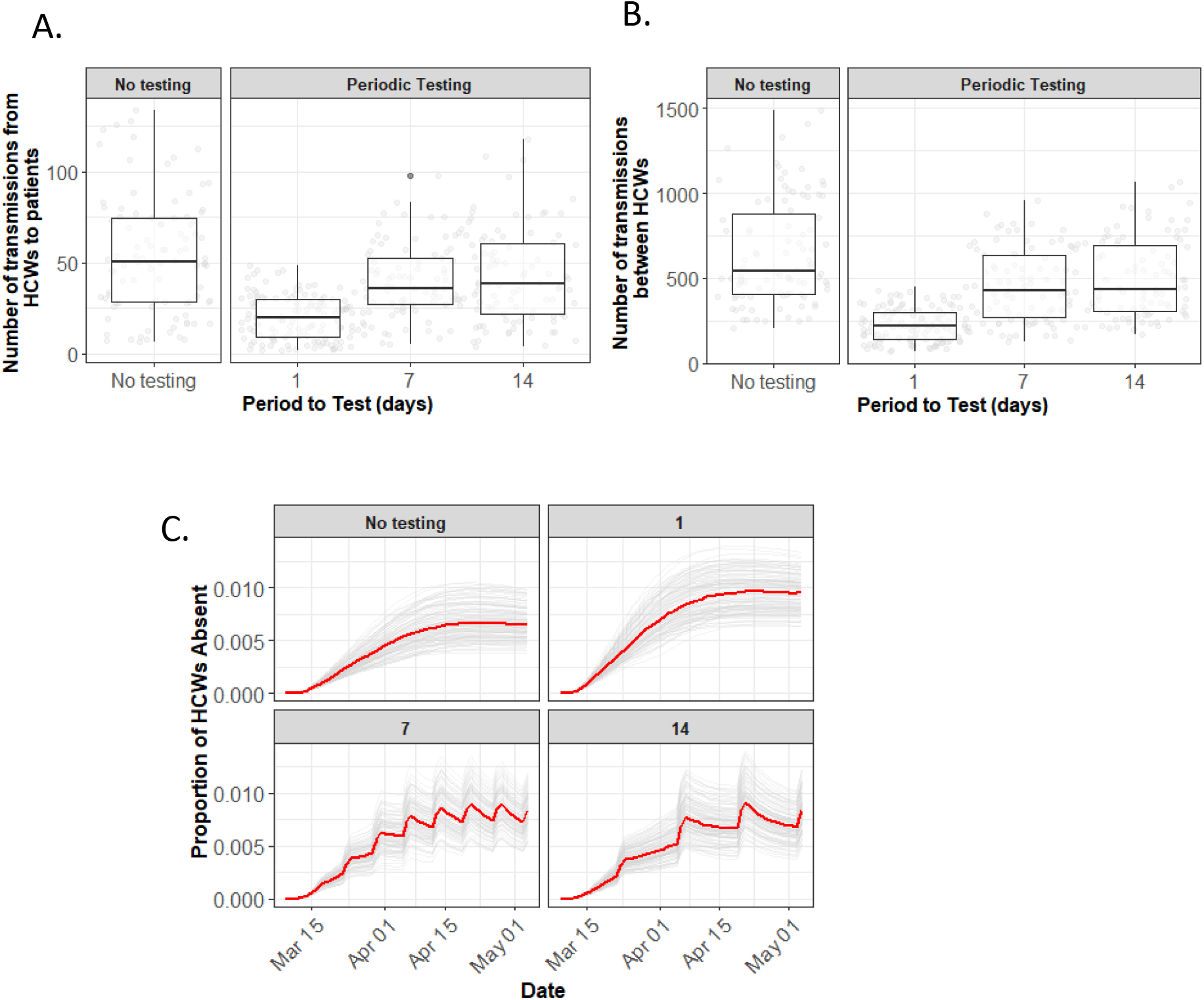
Effect of HCW testing on infections and staffing under Scenario A. A-B) Cumulative number of HCW to patient transmissions (A), and HCW to HCW transmissions (B) between 09-03-2020 - 13-04-2020 under the four different testing strategies. C) Proportion of HCWs absent under each testing strategy (grey lines = individual simulation from 1000 reps, red lines = median).

### Efficacy of single room isolation vs. cohorting of suspected cases on nosocomial transmission events

The majority of nosocomial infections under Scenario B were caused by (direct or indirect) patient to patient transmission (Figure 2C). This suggests that improved infection prevention and control mechanisms within hospitals would be effective in lowering the rate of nosocomial infections in the inpatient population, which would subsequently reduce the number of HCWs infected.

To explore this hypothesis, we simulated a situation where instead of cohorting suspected-positive patients together, suspected patients were isolated in single rooms or bays within a hospital. We assumed that single room isolation reduced the patient to patient transmission rate by 50, 75, and 100%, and that it was feasible for the hospital to place all suspected patients in such single rooms or bays. Under all efficacies, there were significantly fewer nosocomial infections under Scenario B (placing suspected cases in single rooms/bays) than Scenario A (group cohorting of suspected cases)(Figure 4B). In the event that single room isolation in Scenario B was 50% effective at reducing patient to patient transmission events, there were 40% (IQR 13.6, 41.8) fewer nosocomial infections over the simulated time-period, at 75% there were 52% (IQR 36.3, 60.0) fewer nosocomial infections, and at 100% efficacy this reduction was as high as 80% (IQR 56.2, 85.4) (Figure 4B).

## Discussion

Using an SEIR model of transmission, we studied the potential sources of SARS-CoV2 infections in a simulated English hospital. Our results suggest that while the majority of infections in hospital patients are a result of community acquisition of the virus, direct and indirect patient to patient transmission drives nosocomial infection. Furthermore, the majority of cases in HCW were not found to be community acquired, but instead driven by nosocomial transmission. This is in contrast to a recent study that suggests that the rate of asymptomatic infection among HCWs more likely reflects general community transmission than in-hospital exposure (Treibel et al., 2020), although the proportion of infected cases observed by Treibel *et al* is in line with estimates presented in this study. Through the NGM approach of Diekmann et al., (2010) we estimate that the within hospital R0 is 0.45, suggesting that the epidemic within hospitals is not self-sustaining and would die out within four generations without infectious importation. Both patients and other HCWs play an important role in HCW infections (Figure 2).

Given that HCW to HCW infections were identified as a primary contributor to nosocomial infections in the HCW population, we evaluated the potential for periodic testing to reduce infection. In our simulated hospital, we considered a scenario where the entire workforce was tested every 1, 7, or 14 days, and a positive HCW result leading to isolation at home for 7 days from the date of the test (Figure 3). We observed only a small proportion of HCWs absent at any one time (up to 1%) due to testing positive, which is considerably fewer than current observations that are in the region of 25% of staff absent due to suspected COVID-19 infection or self-isolation (NHS Situation Reports). This would therefore suggest that testing HCWs routinely is unlikely to cause an unsustainable decline in the number of available workers.

We suggest that HCW testing will have a moderate effect on patients (up to 14% fewer infections from HCWs) but a larger effect on HCWs (up to 65% fewer HCW to HCW transmissions, and 31% fewer HCW infections in total). This is supported by findings by (Grassly et al., 2020), who suggest that regular screening can prevent transmissions in hospitals and other care facilities.

We find IPC measures, specifically patient isolation, in hospitals to have a larger effect of up to an 80% reduction in nosocomial transmission events (Figure 4).

**Figure 4:**
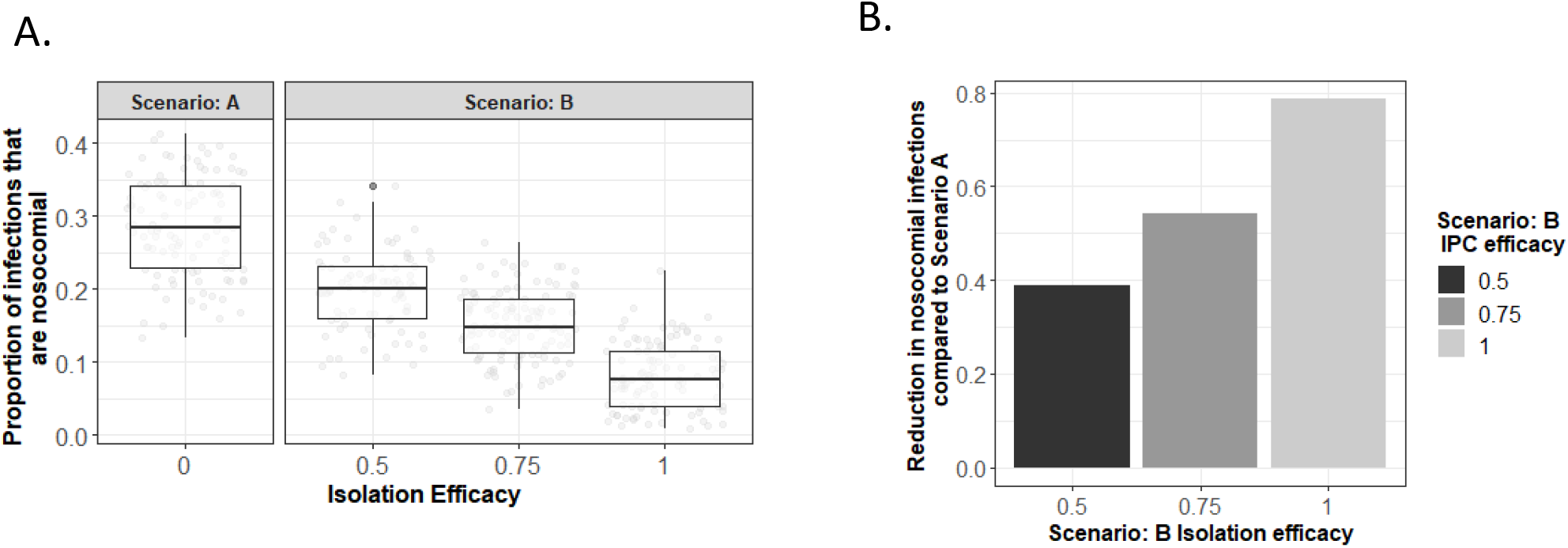
Effect of cohorting and single room isolation with varied isolation efficacy on nosocomial infections. A) Cumulative proportion of infections that are nosocomial between 09-03-2020 and 01-05-2020 under the two isolation scenarios with varying efficacy of single room isolation B) Median reduction in the number of nosocomial infections over varying efficacies of scenario B.

The management of suspected cases on admission to hospital has the potential to dramatically reduce the burden of nosocomial infection. Our model suggests that managing symptomatic patients in single rooms or bays that are fully disinfected in between patients could reduce nosocomial infection rates by up to 80%. Although in practice as the number of community cases increases or the criteria for those patients to be tested changes, it may not be possible to use single rooms or bays, adherence to best practice early in an epidemic can limit the development of further infections and their subsequent effects on community transmission (McDonald et al., 2019; Moloney et al., 2016). Further, this result has important implications when considering the design of temporary facilities such as the newly developed Nightingale Hospitals (NHS England).

While the model is parameterised using the best data currently available, data on contact patterns of (different groups) of HCW with COVID-19 patients and positive beds to determine HCW at risk and HCW infection rates would improve parameterisation and increase certainty in outputs. A limitation of this work is that this model does not separate asymptomatic from pre-symptomatic. Further, the model is deterministic and cannot account for the impact of outbreaks on individual wards, or the sequestering of patients and HCWs onto “hot” and “cold” wards.

Despite these limitations, the results from this work have the potential to impact infection control during coronavirus outbreaks.

## Data Availability

Model code and parameters included in supplement, calibration data privately held.

---

**Figure A1:**
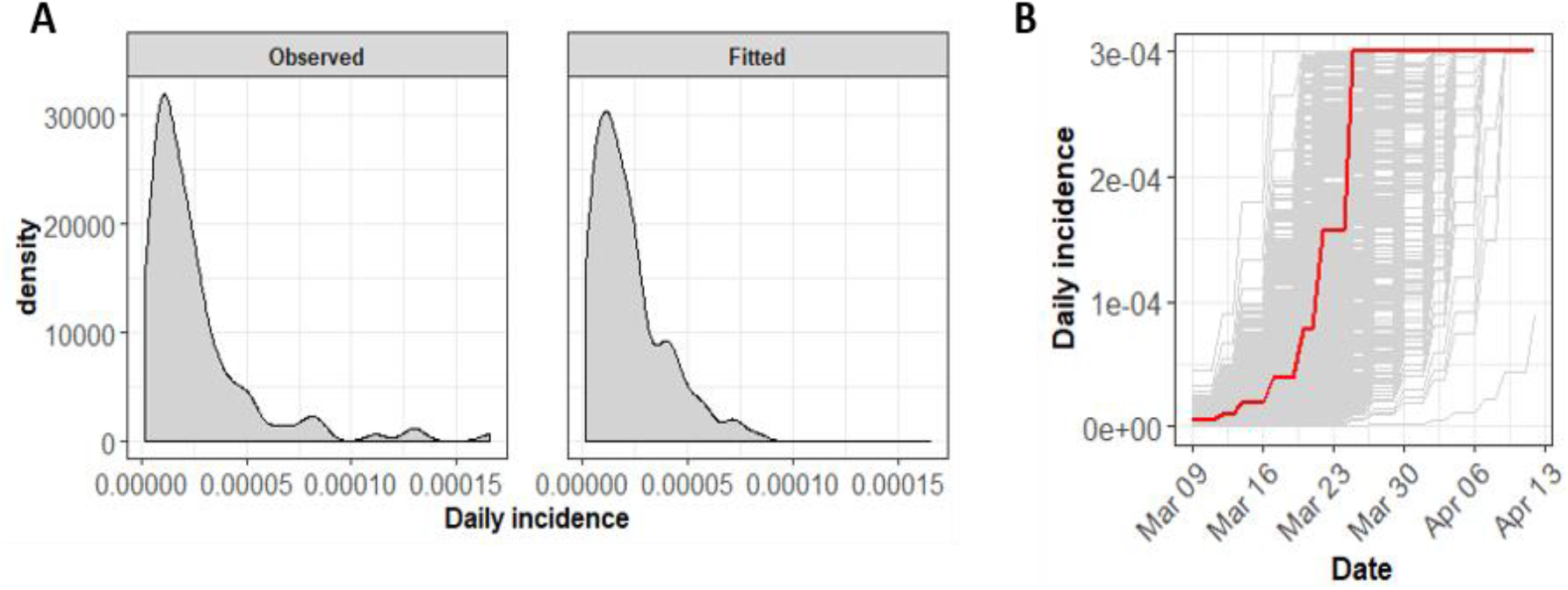
Daily incidence rates. A) Observed and fitted daily incidence rates on 09-03-2020. B) Simulated daily incidence over time in each from 1000 simulations (grey) with median incidence over time (red).

**Figure A2:**
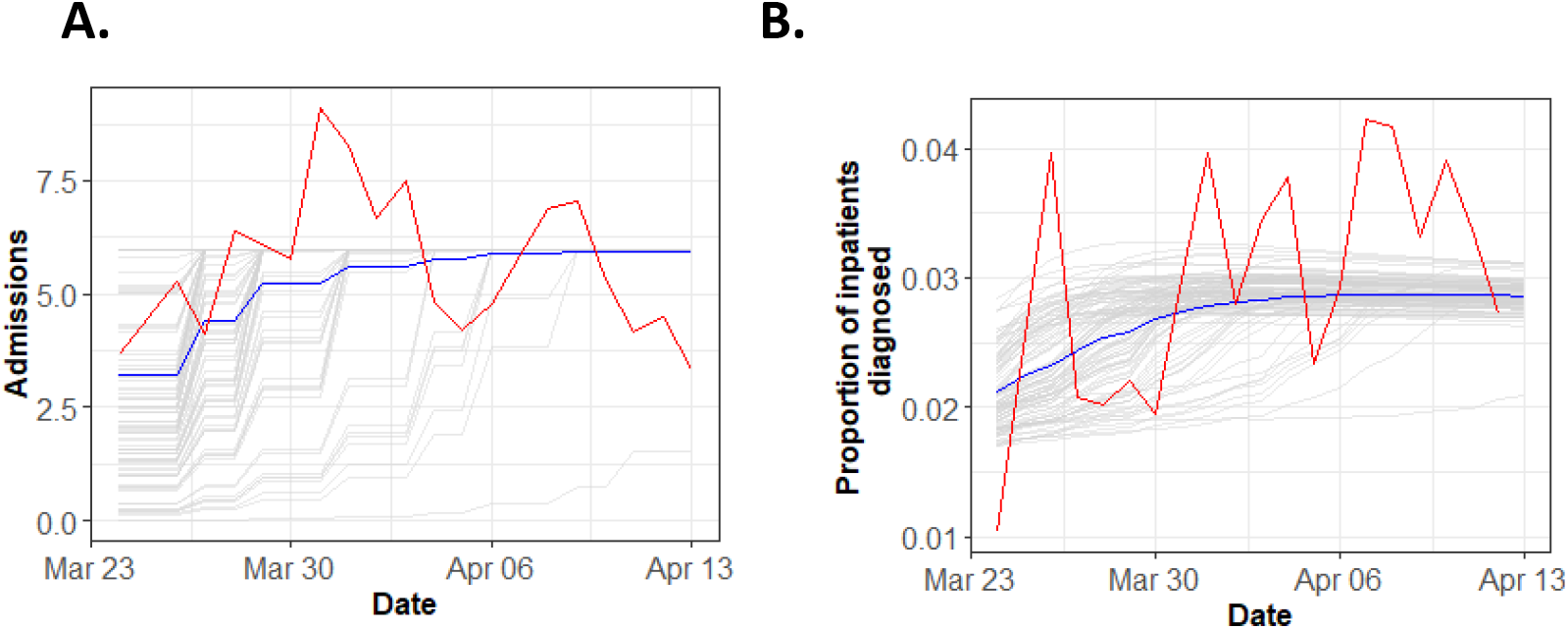
Comparison to NHS Situation Report data: Simulated (blue) and NHS situation report (red) daily admissions data (A) and daily proportion of inpatients developing an infection (B). Grey lines are 100 realisations of simulation.

**Figure A3:**
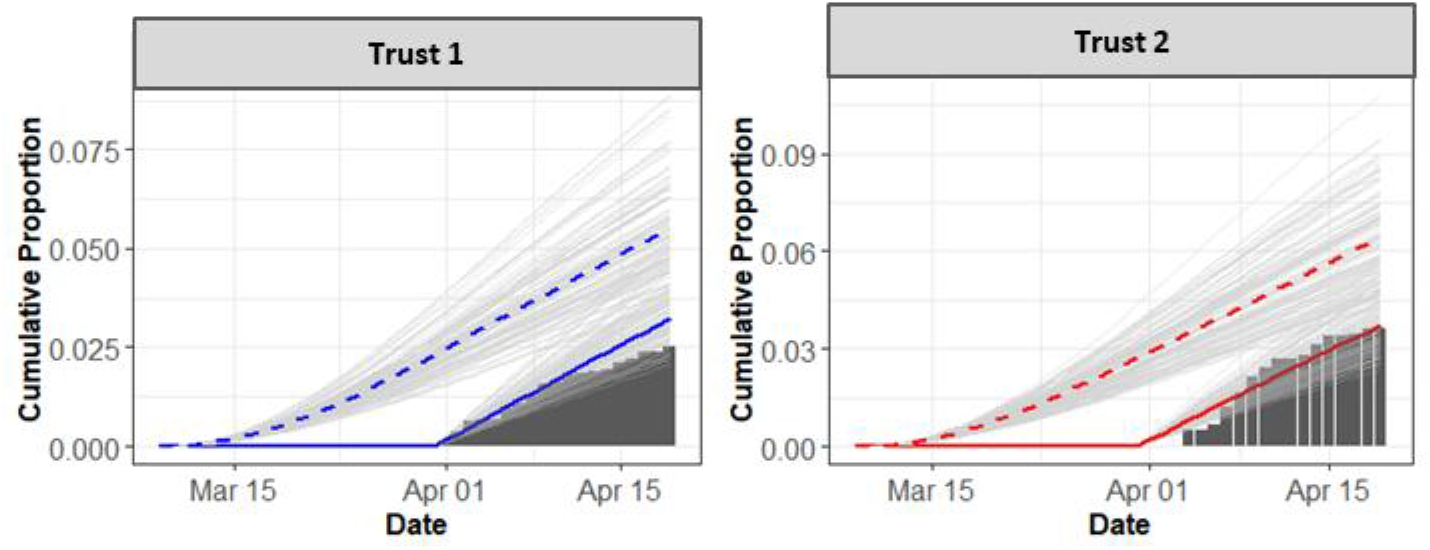
Comparison to swabbing data of symptomatic HCWs from two English Trusts. Simulated symptomatically infected HCWs are shown over swabbing period (solid lines) and total from start of simulation (dashed line).

**Figure A4:**
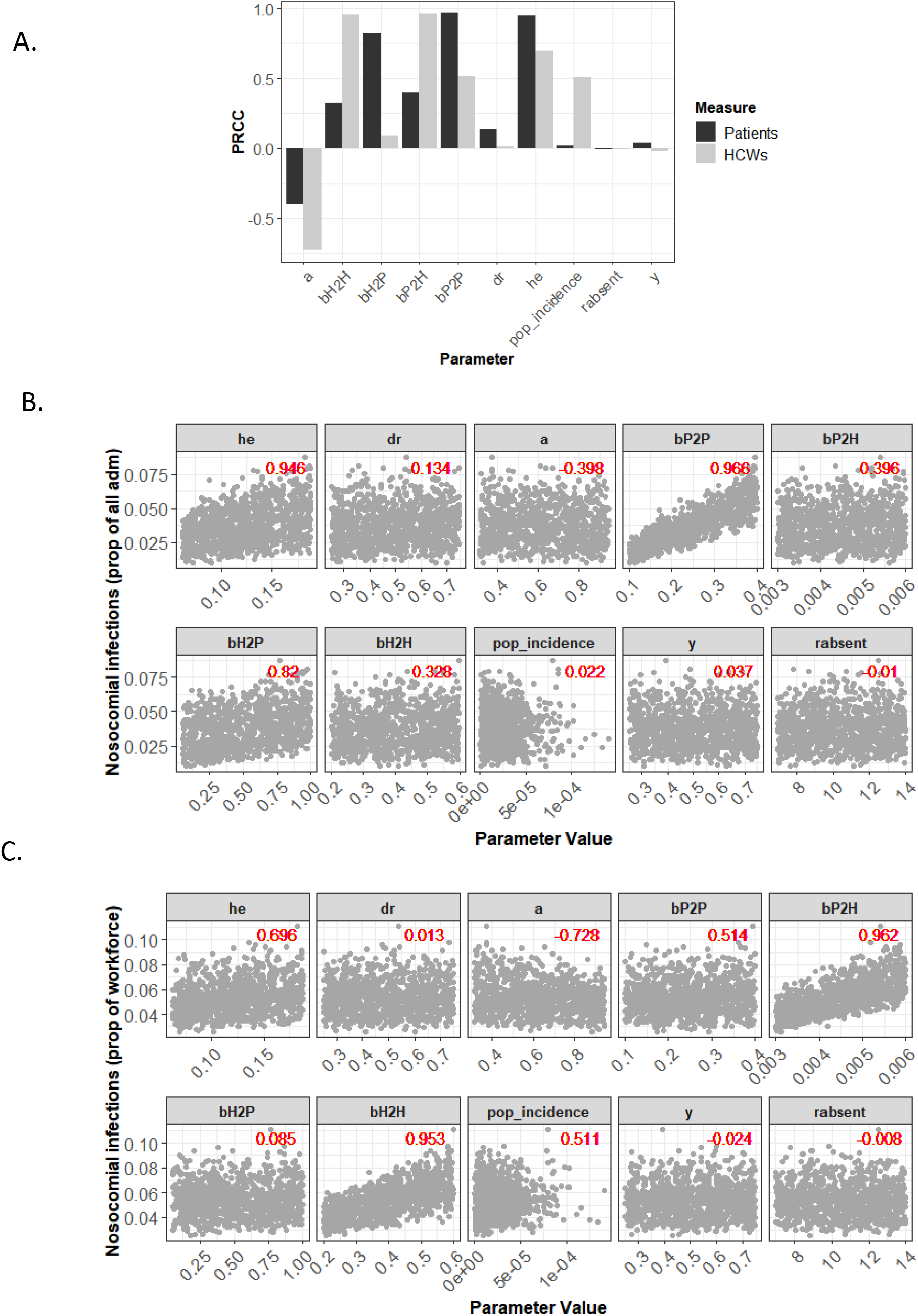
Sensitivity analysis of unknown parameters. A) Partial Rank Correlation Coefficients (PRCCs) for all parameters included in the analysis against the cumulative proportion of nosocomial infections in patients and HCWs. B,C) Parameter values against observed simulation outcome for nosocomial infections in the patient population (B) and HCW population (C). Red text indicates PRCC for each parameter.

## References

Alden, K., Read, M., Timmis, J., Andrews, P.S., Veiga-Fernandes, H., and Coles, M. (2013). Spartan: A Comprehensive Tool for Understanding Uncertainty in Simulations of Biological Systems. PLOS Computational Biology 9, e1002916.

Bin, S.Y., Heo, J.Y., Song, M.-S., Lee, J., Kim, E.-H., Park, S.-J., Kwon, H., Kim, S. mi, Kim, Y., Si, Y.-J., et al. (2016). Environmental Contamination and Viral Shedding in MERS Patients During MERS-CoV Outbreak in South Korea. Clin Infect Dis 62, 755–760.

Chowell, G., Abdirizak, F., Lee, S., Lee, J., Jung, E., Nishiura, H., and Viboud, C. (2015). Transmission characteristics of MERS and SARS in the healthcare setting: a comparative study. BMC Med 13.

Daskalaki, I., Hennessey, P., Hubler, R., and Long, S.S. (2007). Resource consumption in the infection control management of pertussis exposure among healthcare workers in pediatrics. Infect Control Hosp Epidemiol 28, 412–417.

Diekmann, O., Heesterbeek, J.A.P., and Roberts, M.G. (2010). The construction of next-generation matrices for compartmental epidemic models. J R Soc Interface 7, 873–885.

Dixon, M.G., Taylor, M.M., Dee, J., Hakim, A., Cantey, P., Lim, T., Bah, H., Camara, S.M., Ndongmo, C.B., Togba, M., et al. (2015). Contact Tracing Activities during the Ebola Virus Disease Epidemic in Kindia and Faranah, Guinea, 2014. Emerg Infect Dis 21, 2022–2028.

England, D.O., Currie, M.J., and Bowden, F.J. (2005). An audit of contact tracing for cases of chlamydia in the Australian Capital Territory. Sex Health 2, 255–258.

Grad, Y.H., and Lipsitch, M. (2014). Epidemiologic data and pathogen genome sequences: a powerful synergy for public health. Genome Biol 15, 1–14.

Grassly, N.C., and Fraser, C. (2008). Mathematical models of infectious disease transmission. Nat. Rev. Microbiol. 6, 477–487.

Grassly, N.C., Pons-Salort, M., Parker, E.P., White, P.J., Ainslie, K., Baguelin, M., Bhatt, S., Blake, I., Boonyasiri, A., Boyd, O., et al. (2020). Report 16: Role of testing in COIVD-19 control. 13.

Houlihan, C.F., Frampton, D., Ferns, R.B., Raffle, J., Grant, P., Reidy, M., Hail, L., Thomson, K., Mattes, F., Kozlakidis, Z., et al. (2018). Use of Whole-Genome Sequencing in the Investigation of a Nosocomial Influenza Virus Outbreak. J. Infect. Dis. 218, 1485–1489.

Hunter, E., Price, D.A., Murphy, E., Loeff, I.S. van der Baker, K.F., Lendrem, D., Lendrem, C., Schmid, M.L., Pareja-Cebrian, L., Welch, A., et al. (2020). First experience of COVID-19 screening of health-care workers in England. The Lancet 0.

Hunter, J.C., Nguyen, D., Aden, B., Al Bandar, Z., Al Dhaheri, W., Abu Elkheir, K., Khudair, A., AI Mulla, M., El Saleh, F., Imambaccus, H., et al. (2016). Transmission of Middle East Respiratory Syndrome Coronavirus Infections in Healthcare Settings, Abu Dhabi. Emerging Infect. Dis. 22, 647–656.

Keeley, A.J., Evans, C., Colton, H., Ankcorn, M., Cope, A., State, A., Bennett, T., Giri, P., Silva, T.I. de, and Raza, M. (2020). Roll-out of SARS-CoV-2 testing for healthcare workers at a large NHS Foundation Trust in the United Kingdom, March 2020. Eurosurveillance 25, 2000433.

Keeling, M.J., and Danon, L. (2009). Mathematical modelling of infectious diseases. Br. Med. Bull. 92, 33–42.

Kluytmans, M., Buiting, A., Pas, S., Bentvelsen, R., Bijllaardt, W. van den Oudheusden, A. van Rijen, M. van Verweij, J., Koopmans, M., and Kluytmans, J. (2020). SARS-CoV-2 infection in 86 healthcare workers in two Dutch hospitals in March 2020. MedRxiv 2020.03.23.20041913.

Linton, N.M., Kobayashi, T., Yang, Y., Hayashi, K., Akhmetzhanov, A.R., Jung, S.-M., Yuan, B., Kinoshita, R., and Nishiura, H. (2020). Incubation Period and Other Epidemiological Characteristics of 2019 Novel Coronavirus Infections with Right Truncation: A Statistical Analysis of Publicly Available Case Data. J Clin Med 9.

McDonald, E.G., Dendukuri, N., Frenette, C., and Lee, T.C. (2019). Time-Series Analysis of Health Care-Associated Infections in a New Hospital With All Private Rooms. JAMA Intern Med.

Moloney, G., Mac Aogáin, M., Kelleghan, M., O’Connell, B., Hurley, C., Montague, E., Conlon, M., Murray, H., and Rogers, T.R. (2016). Possible Interplay Between Hospital and Community Transmission of a Novel Clostridium Difficile Sequence Type 295 Recognized by Next-Generation Sequencing. Infect Control Hosp Epidemiol 37, 680–684.

NHS England NHS England » NHS steps up coronavirus fight with two more Nightingale Hospitals.

Pung, R., Chiew, C.J., Young, B.E., Chin, S., Chen, M.I.-C., Clapham, H.E., Cook, A.R., Maurer-Stroh, S., Toh, M.P.H.S., Poh, C., et al. (2020). Investigation of three clusters of COVID-19 in Singapore: implications for surveillance and response measures. The Lancet 395, 1039–1046.

Sanche, S., Lin, Y.T., Xu, C., Romero-Severson, E., Hengartner, N., and Ke, R. Early Release - High Contagiousness and Rapid Spread of Severe Acute Respiratory Syndrome Coronavirus 2 - Volume 26, Number 7–July 2020 - Emerging Infectious Diseases journal - CDC.

Tindale, L., Coombe, M., Stockdale, J.E., Garlock, E., Lau, W.Y.V., Saraswat, M., Lee, Y.-H.B., Zhang, L., Chen, D., Wallinga, J., et al. (2020). Transmission interval estimates suggest pre-symptomatic spread of COVID-19. MedRxiv 2020.03.03.20029983.

Treibel, T.A., Manisty, C., Burton, M., McKnight, Á., Lambourne, J., Augusto, J.B., Couto-Parada, X., Cutino-Moguel, T., Noursadeghi, M., and Moon, J.C. (2020). COVID-19: PCR screening of asymptomatic health-care workers at London hospital. The Lancet 0.

Wang, D. (2020). Clinical Characteristics of 138 Hospitalized Patients With 2019 Novel Coronavirus-Infected Pneumonia in Wuhan, China. - PubMed - NCBI.

Wang, X., Pan, Z., and Cheng, Z. (2020). Association between 2019-nCoV transmission and N95 respirator use. Journal of Hospital Infection 0.

de Wit, E., van Doremalen, N., Falzarano, D., and Munster, V.J. (2016). SARS and MERS: recent insights into emerging coronaviruses. Nat Rev Microbiol 14, 523–534.

Zhou, Q., Gao, Y., Wang, X., Liu, R., Du, P., Wang, X., Zhang, X., Lu, S., Wang, Z., Shi, Q., et al. (2020). Nosocomial Infections Among Patients with COVID-19, SARS and MERS: A Rapid Review and Meta-Analysis. MedRxiv 2020.04.14.20065730.

